# Spine age estimation using deep learning in lateral spine radiographs and DXA VFA to predict incident fracture and mortality

**DOI:** 10.1101/2025.03.25.25324645

**Authors:** Sang Wouk Cho, Namki Hong, Kyoung Min Kim, Young Han Lee, Chang Oh Kim, Hyeon Chang Kim, Yumie Rhee, Brian H. Chen, William D. Leslie, Steven R. Cummings

## Abstract

**Background:** Spine age estimated from lateral spine radiographs and DXA vertebral fracture assessments (VFAs) could be associated with fracture and mortality risk.

**Methods:** In the VERTE-X cohort (n=10,341, age 40 or older; derivation set) and KURE cohort (n=3,517; age 65 or older; external test set), predicted age difference was defined as estimated spine age minus chronological age. The primary outcome was incident fracture. Secondary outcomes included morphologic vertebral fracture, osteoporosis, and incident mortality.

**Results:** Incidence of overall fracture was 20.5/1000 and 21.0/1000 person-years (median follow-up 5.4 and 6.6 years) in VERTE-X and KURE, respectively. Spine age discriminated prevalent vertebral fractures and osteoporosis better than chronological age. Higher predicted age difference (PAD) was associated with greater risk of overall (VERTE-x: adjusted HR [aHR] 1.71; KURE: aHR 1.22 per 1 standard deviation [SD] increment), vertebral (aHR 1.55 and 1.34), and non-vertebral fractures (aHR 1.89 and 1.15, p<0.05 for all), independent of chronological age and prevalent vertebral fracture. FRAX hip fracture probabilities based on spine age improved discrimination for incident hip fracture over chronological age (AUROC 0.83 vs. 0.78, p=0.027). Shorter height, lower femoral neck BMD, diabetes, vertebral fractures, and surgical prosthesis were associated with higher predicted age difference, explaining 40% of variance. In the external test set, higher predicted age difference was associated with greater risk of mortality (aHR 1.31 per 1 SD increment, p=0.001), independent of covariates.

**Conclusion:** Spine age estimated from lateral spine radiographs and DXA VFA enhanced fracture risk assessment and mortality prediction in adults.

**Key Points:**

- Spine age estimated from lateral spine radiographs and DXA VFA using deep learning outperformed chronological age in discriminating morphologic vertebral fracture and osteoporosis.
- Higher predicted age difference (predicted spine age minus chronological age) was associated with greater risk of overall, vertebral, and non-vertebral incident fracture, independent of covariates.
- Male sex, lower height, lower femoral neck BMD, diabetes mellitus, morphologic vertebral fractures, and surgical prosthesis were correlated with higher predicted age difference, explaining up to 40% variance.
- Higher predicted age difference was associated with greater risk of mortality, independent of chronological age, sex, prevalent morphologic vertebral, fracture, and clinical biomarkers related to mortality including serum albumin, hemoglobin, and creatinine.

## Introduction

Fractures are a major health burden related to aging, posing significant morbidity and mortality globally.^1^ According to the Global Burden of Disease study, the absolute incidence, years lived with disability, and health care costs for fracture increased substantially between 1990 and 2019, with highest incidence in the oldest old age group.^1^ Although several fracture risk assessment tools are available, there is room to improve performance of individualized assessment to facilitate the optimal use of pharmacologic interventions for fracture prevention.^2^

Biological age represents the state of the body or organ system of an individual estimated as an integrated value of biophysiological measures in contrast to the chronological age as the time since the individual’s birth.^3^ Biological age, estimated from various imaging modalities including brain magnetic resonance images (MRI),^4^ eye retinal photographs,^5^ and chest radiographs,^6^ outperformed chronological age in predicting health outcomes including mortality. However, attempts to estimate biological age in the musculoskeletal system using imaging data are limited. If the estimation of biological age to accurately reflect musculoskeletal status is feasible using simple and widely available image sources, such as spine radiographs, then it could improve individual fracture risk assessment over chronological age in clinical practice.

In this study, we developed a convolutional neural network model to estimate spine age from lateral spine radiographs and dual energy X-ray absorptiometry (DXA) vertebral fracture assessment (VFA) images. Discriminatory performance for prevalent vertebral fracture and osteoporosis was compared for biological spine age versus chronological age. Prognostic value of predicted spine age difference for incident fracture and mortality was assessed with adjustment for chronological age, sex, and covariates.

## Materials and methods

### Study participants

Derivation cohort: Demographic and clinical data of individuals who underwent lateral spine radiographs at Severance Hospital, Seoul, Korea between January 2007 and December 2018 were collected (Supplementary Figure 1; the VERTEbral fracture and osteoporosis detection in spine X-ray study, VERTE-X).^8^ Requirement for written permission for the medical record review was waived by Institutional Review Board (IRB no. 4-2021-0937). Cohort entry date (index date) was the date of initial spine radiograph acquisition. We excluded individuals with age younger than 40 years, history of bone metastasis or hematologic malignancy within 1 year prior to index date, severe scoliosis, kyphosis, poor image quality, non-Korean ethnicity, and those without follow-up radiographs at least 28 days after the index date. A total of 10,341 participants remained in the final cohort. The dataset was randomly split into train (60%), validation (20%), and a hold-out test set (20%).

External test cohort: Korean Urban Rural Elderly (KURE) cohort is a prospective cohort study of aging and health outcomes in community-dwelling older adults.^19^ Details on the cohort have previously been published.^19,20^ A total of 3,517 individuals aged 65 years or older participated in the study at baseline (year 2012-2015) after obtaining written permission (IRB no 4-2012-0172). After excluding individuals without DXA VFA images (n=9), a total of 3,508 participants remained in the external test cohort.

### Spine age estimation

Model architecture for the convolutional neural network (CNN) to estimate spine age is presented in Supplementary Figure 2. Using the EfficientNet-B4 architecture, we applied the mean-variance loss function proposed by Pan and colleagues.^21^ This approach simultaneously penalizes the difference between the mean of the predicted spine age distribution and the chronological age (mean loss) and reduces the variance of the estimated distribution to maintain a concentrated prediction (variance loss). Although lateral spine radiographs and VFA images differ in their resolution, they share similar positioning and morphological features. Thus, we were able to extract features and predict spine age using a single CNN model for both modalities. To reduce modality-specific differences, an age-level bias correction was applied to the external VFA test set based on Beheshti’s method^22,23^. Further details on model hyperparameters are described in the supplementary method. To enhance model explainability, we visualized regions of high importance in spine age prediction using gradient-weighted class activation mapping (Grad-CAM), which highlighted specific regions with strong weights from the CNN kernel.^24^ All results from the spine age prediction model and Grad-CAM visualizations are available at https://bonecentriq.org.

### Outcomes

In the derivation set (lateral spine radiographs, VERTE-X), incident fracture was defined as any new-onset morphologic vertebral fracture confirmed on follow-up lateral spine radiographs or clinical non-vertebral fractures at the hip, distal forearm, upper arm, pelvis, and lower leg ascertained during follow-up from Severance Hospital electronic medical records system until the last observation date (December 31, 2023). In the external test cohort (DXA VFA, KURE) of community-dwelling older adults, outcomes including clinical fracture (any clinical vertebra, hip, distal forearm, or proximal humerus fracture) and mortality were collected by interviewer-assisted questionnaire performed at the time of four-year interval follow-up visit to the center or follow-up phone calls until December 31^st^, 2021, with subsequent outcome ascertainment using individual-level diagnosis codes and/or procedural codes obtained by linkage to the Health Insurance Review and Assessment (HIRA; research data [M20190729878]) which covers 99% of residents in South Korea.^25^

### Covariates

Information on chronological age, sex, height, weight, previous history of clinical fracture, chronic glucocorticoid use, and presence of rheumatoid arthritis at the time of cohort entry (index date) were collected by reviewing electronic health record of Severance Hospital in the derivation set (VERTE-X). In the external test set (KURE), information on covariates were collected at the time of cohort entry using interviewer-assisted questionnaires and anthropometry measurements. FRAX probability (Korean tool) for major osteoporotic fracture and hip fracture without BMD were calculated using an online calculator (https://frax.shef.ac.uk/frax/tool.aspx?country=25; web version 1.4.7). Unavailable covariates to calculate FRAX were entered as ‘no’ responses. High risk FRAX probabilities were defined as FRAX major osteoporotic fracture probability ≥ 20% or FRAX hip fracture probability ≥ 3%.^26^ DXA areal bone mineral density (BMD) measurement (Discovery W and A, Hologic, USA) was available in 70% (1448/2063) of the derivation test set (VERTE-X) and 100% (3508/3508) of the external test set (KURE). Presence of osteoporosis was defined as DXA areal BMD T-score −2.5 or below (reference population: NHANES III young White female) from the lumbar spine, femoral neck, or total hip.^27^

### Statistical analysis

Differences in clinical characteristics of study participants with or without incident fracture outcomes were compared using two-sample independent t-tests for continuous variables (presented as mean ± standard deviation) and chi-square test for categorical variables (presented as number and percentage). Discriminatory ability for the presence of morphologic vertebral fracture and osteoporosis at baseline was compared between chronological age and predicted spine age using the area under the receiver-operating characteristics (AUROC) by the De Long method.^28^ Kaplan-Meier failure curves were plotted for incident fracture grouped by chronological age and predicted age difference (predicted spine age minus chronological age) among participants categorized as four groups based upon chronological age (above or below median) and presence of accelerated spine age (defined as highest tertile of predicted age difference [spine age minus chronological age] in the derivation test set, +1 year or higher). Proportional Cox hazard models were built to assess the association between predicted age difference (per one standard deviation increment) and incident fracture, with adjustments for age, sex, presence of prevalent vertebral fracture, clinical risk factors and osteoporosis.

## Results

### Characteristics of study participants

Median follow-up duration was 5.4 years (interquartile range [IQR] 2.6 to 7.7 years) in the derivation test set (VERTE-X) and 6.6 years (IQR 5.5 to 7.5 years) in the external test set (KURE). Incidence rates of overall fracture events during follow-up were 20.5/1000 person-years and 21.0/1000 person-years in the derivation test set (251/2063, 12.2%) and external test set (473/3508, 13.5%), respectively. Participants with versus without incident fracture during follow-up had older chronological and predicted spine age, with a higher prevalence of women, previous history of fracture, morphometric vertebral fracture, surgical prosthesis in the spine, and lower DXA areal BMD.

### Discriminatory ability of chronological age and predicted spine age

The Pearson correlation coefficient between chronological age and spine age was 0.88 and 0.50 in the derivation test set and external test set (both p<0.001), respectively (Supplementary Figure 3). Average predicted age difference (spine age minus chronological age) was −0.8 years (standard deviation 4.9) in the derivation test set and −0.5 years (standard deviation 8.0) in the external test set. Spine age showed better discriminatory performance for the presence of morphologic vertebral fracture or osteoporosis in both the derivation test set (Figure 1; AUROC: vertebral fracture, 0.77 vs. 0.72; osteoporosis, 0.66 vs. 0.52) and external test set (AUROC: vertebral fracture, 0.66 vs. 0.60; osteoporosis, 0.65 vs. 0.57, all p<0.001).

**Figure 1.**
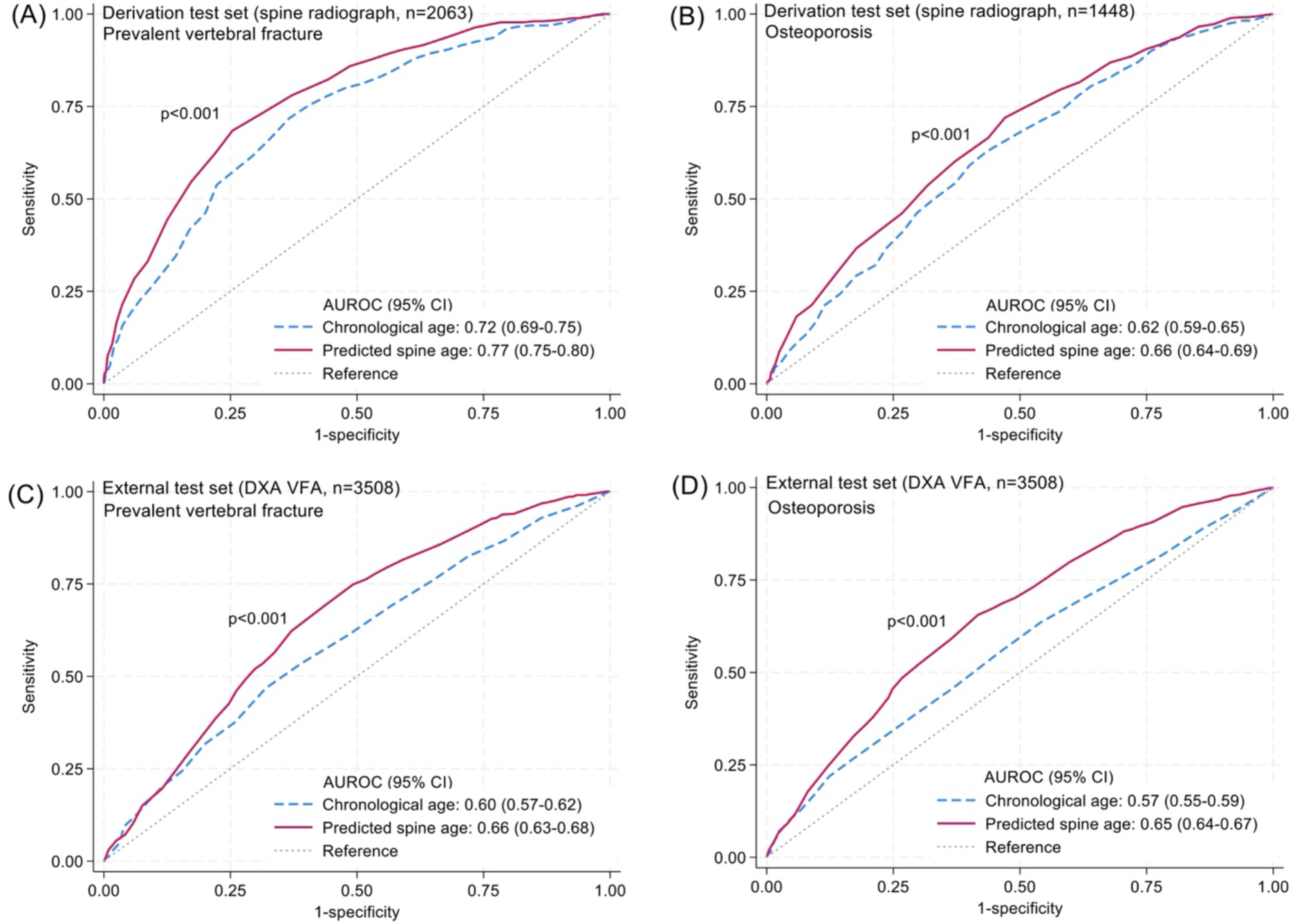
Discriminatory ability of chronological age and predicted spine age for prevalent vertebral fracture (A and C) and osteoporosis (B and D)

### Association of predicted age differences with incident fracture

In Figure 2, compared to the referent group of individuals of younger chronological age (below median; 65 years in the derivation test set and 72 years in the external test set) without accelerated spine age, younger chronological age with accelerated spine age was associated with a 2.21- and 1.60-fold elevated fracture risk in the derivation and external test sets, respectively. Participants of older chronological age (above the median) with accelerated spine age had the highest fracture risk in both cohorts (unadjusted hazard ratio [HR] 6.67 and 2.53 in derivation and external test sets, respectively). In the derivation and external test sets, each standard deviation increment of predicted age difference was associated with greater risk of overall (adjusted HR [aHR] 1.71 and 1.22, respectively), vertebral (aHR 1.55 and 1.34), and non-vertebral fractures (aHR 1.89 and 1.15, all p<0.05), independent of age, sex, prevalent morphologic vertebral fracture, clinical risk factors, and osteoporosis (Table 2).

**Figure 2.**
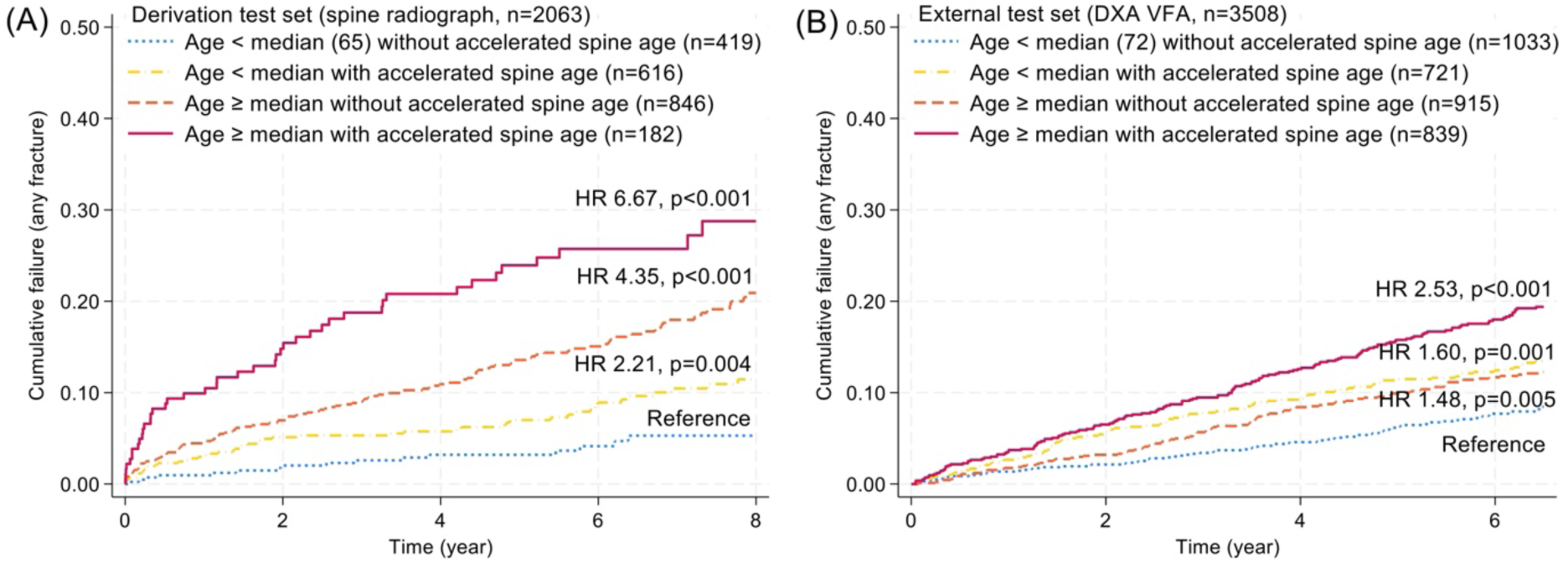
Kaplan-Meier cumulative failure curve for overall incident clinical fracture in the (A) spine radiograph cohort (derivation test set) and (B) DXA VFA cohort (external test set) from combinations of chronological age (< or ≥ median) and predicted spine age (accelerated spine age versus non-accelerated age). Accelerated spine age was defined using highest tertile threshold of predicted age difference (predicted spine age minus chronological age ≥ +1) in the derivation test set. Abbreviations: HR: unadjusted hazard ratio.

**Table 1.** Clinical characteristics of study participants.

|  | Lateral spine radiograph cohort (VERTE-X, hospital-based) age 40 years or older; derivation test set (n=2063) |  |  | DXA VFA cohort (KURE, community-based) age 65 years or older; external test set (n=3508) |  |  |
| --- | --- | --- | --- | --- | --- | --- |
|  | Incident fracture during follow-up |  |  | Incident fracture during follow-up |  |  |
|  | No (n=1812) | Yes (n=251) | P-value | No (n=3035) | Yes (n=473) | P-value |
| Age, years | 63.0 ± 10.2 | 68.5 ± 8.4 | <0.001 | 71.7 ± 4.6 | 73.1 ± 4.8 | <0.001 |
| Predicted spine age, years | 62.1 ± 8.4 | 67.7 ± 6.3 | <0.001 | 70.8 ± 9.3 | 74.8 ± 8.6 | <0.001 |
| Women, n (%) | 1127 (62.2) | 200 (79.7) | <0.001 | 1969 (64.9) | 380 (80.3) | <0.001 |
| Body mass index, kg/m<sup>2</sup> | 24.2 ± 3.3 | 23.8 ± 3.4 | 0.075 | 24.2 ± 3.0 | 24.1 ± 3.0 | 0.346 |
| Previous clinical fracture, n (%) | 88 (4.9) | 55 (21.9) | <0.001 | 439 (20.7) | 90 (26.9) | 0.011 |
| Morphologic vertebral fracture, n (%)** | 265 (14.6) | 90 (35.9) | <0.001 | 337 (11.1) | 97 (20.5) | <0.001 |
| Surgical prosthesis in spine, n (%) | 220 (12.1) | 44 (17.5) | 0.017 | 98 (3.2) | 25 (5.3) | 0.024 |
| Chronic glucocorticoid use, n (%) | 74 (4.1) | 26 (10.4) | <0.001 | 8 (0.3) | 1 (0.2) | 0.835 |
| Rheumatoid arthritis, n (%) | 47 (2.6) | 17 (6.8) | <0.001 | 29 (1.0) | 6 (1.3) | 0.524 |
| DXA aBMD T-score* |  |  |  |  |  |  |
| Lumbar spine | -1.2 ± 1.9 | -2.3 ± 1.7 | <0.001 | -1.6 ± 1.6 | -2.4 ± 1.4 | <0.001 |
| Femoral neck | -1.6 ± 1.2 | -2.3 ± 1.1 | <0.001 | -1.9 ± 0.9 | -2.4 ± 0.9 | <0.001 |
| Total hip | -1.1 ± 1.2 | -1.9 ± 1.1 | <0.001 | -1.3 ± 1.0 | -1.8 ± 0.9 | <0.001 |
| Osteoporosis, n (%)# | 452 (35.0) | 97 (62.2) | <0.001 | 1272 (41.9) | 306 (64.7) | <0.001 |

\*Available in 1,448/2063 in VERTE-X dataset. †FRAX probability without BMD. \*\*Morphologic vertebral fracture was defined according to modified algorithm-based qualitative (mABQ) method. #Osteoporosis was defined as DXA areal BMD T-score -2.5 or below at lumbar, femoral neck, or total hip using NHANES III White young female reference. Abbreviations: DXA, dual-energy X-ray absorptiometry; aBMD, areal bone mineral density; VFA, vertebral fracture assessment; VERTE-X, vertebral X-ray cohort; KURE, Korean urban rural elderly cohort.

**Table 2.** Association of predicted age difference with incident fracture outcome in the derivation test set (spine radiograph cohort, n=2063) and external test set (DXA VFA cohort, n=3508)

| Predictor: predicted age difference, per 1 SD increment | Model 1 (age- and sex-adjusted) |  | Model 2 (model 1 + prevalent vertebral fracture and clinical risk factors*) |  | Model 3 (model 2 + osteoporosis**) |  |
| --- | --- | --- | --- | --- | --- | --- |
|  | HR (95% CI) | P-value | HR (95% CI) | P-value | HR (95% CI)<sup>†</sup> | P-value |
| Outcome: incident clinical fracture |  |  |  |  |  |  |
| Derivation test set (lateral spine radiograph, VERTE-X cohort) |  |  |  |  |  |  |
| Overall | 1.74 (1.46 to 2.07) | <0.001 | 1.54 (1.29 to 1.84) | <0.001 | 1.71 (1.35 to 2.18) | <0.001 |
| Vertebral | 1.63 (1.31 to 2.02) | <0.001 | 1.49 (1.19 to 1.86) | <0.001 | 1.55 (1.15 to 2.08) | 0.004 |
| Nonvertebral | 1.92 (1.49 to 2.47) | <0.001 | 1.61 (1.24 to 2.09) | <0.001 | 1.89 (1.31 to 2.71) | 0.001 |
| External test set (DXA VFA, KURE cohort) |  |  |  |  |  |  |
| Overall | 1.28 (1.16 to 1.42) | <0.001 | 1.26 (1.14 to 1.40) | <0.001 | 1.22 (1.10 to 1.35) | <0.001 |
| Vertebral | 1.41 (1.21 to 1.61) | <0.001 | 1.37 (1.19 to 1.59) | <0.001 | 1.34 (1.16 to 1.54) | <0.001 |
| Nonvertebral | 1.20 (1.05 to 1.38) | 0.006 | 1.19 (1.04 to 1.36) | 0.011 | 1.15 (1.01 to 1.32) | 0.042 |

\*Height, previous history of clinical fracture or morphometric vertebral fracture in spine images, surgical prosthesis in spine images, rheumatoid arthritis, and chronic glucocorticoid use. <sup>†</sup>In derivation test set, BMD data were available in a subset (n=1441). \*\*DXA areal BMD T-score -2.5 or lower at lumbar spine, femoral neck, or total hip.

### Reclassification of FRAX risk categories by predicted spine age

When FRAX probabilities for major osteoporotic fracture and hip fracture were calculated using predicted spine age in the place of chronological age in the derivation test set, up-classification from low to high risk group was observed in 53 individuals (53/2063, 2.5%), whereas 144 were down-classified from high to low risk (144/2063, 6.9%; Supplementary Figure 4). Individuals who remained at high risk had the highest fracture risk (28.9%), followed by the low to high up-classified group (24.5%). High risk by FRAX probabilities based on estimated spine age versus FRAX probabilities from chronological age demonstrated higher positive predictive value (28.3% vs. 25.2%, respectively) but lower sensitivity (40.6% vs. 45.4%), yielding modest improvement in odds ratios (4.1 vs. 3.6) for incident fracture (Supplementary Table 1). Using image-predicted spine age instead of chronological age improved the discriminatory performance of FRAX probabilities to predict hip fracture (FRAX MOF probabilities: AUROC 0.81 vs. 0.77, p=0.007; FRAX hip fracture probabilities: 0.83 vs. 0.78, p=0.027; Supplementary Table 2) and FRAX hip fracture probabilities to predict overall fractures (AUROC 0.74 vs. 0.72, p=0.024), although the difference in discriminatory performance for overall incident fracture between FRAX MOF probabilities based on spine age and chronological age to did not reach statistical significance (AUROC 0.74 vs. 0.73, p=0.097).

### Factors associated with predicted age difference

In Supplementary Figure 5, examples of GRAD-CAM (visualizing the pixels with the largest influence on the CNN model’s spine age prediction) for spine radiographs from individuals with and without accelerated spine age are presented. Images with accelerated spine age showed the presence of morphologic vertebral fracture (Supplementary Figure 5A), vertebroplasty or surgical prosthesis (Supplementary Figure 5B), and aortic calcification with various degrees of degenerative changes (Supplementary Figure 5C). Male sex (+0.66 year vs. women), lower height (+0.31 year per 5 cm decrement), presence of diabetes mellitus (+0.51 year), prevalent morphometric vertebral fracture (+1.64 year), lower femoral neck BMD (+0.64 year per 1 standard deviation decrement), and presence of surgical prosthesis (+1.15 year) were associated with higher predicted age difference in a multivariable linear regression model, with about 40% of variance in predicted age difference explained by the model (adjusted R^2^ 0.40; Supplementary Table 3).

### Spine age and mortality

In the external test set of community-dwelling older adults, individuals with accelerated spine age had elevated risk of mortality compared to those without (unadjusted HR 1.36, p=0.036; Supplementary Figure 6). Higher predicted age difference was associated with greater risk of mortality, independent of chronological age, sex, prevalent morphologic vertebral, fracture, and clinical biomarkers related to mortality including serum albumin, hemoglobin, and creatinine (Table 3; adjusted HR 1.31 per 1 standard deviation increment in predicted age difference, 95% CI 1.12 to 1.53, p=0.001).

**Table 3.** Association of predicted spine age difference using DXA VFA with all-cause mortality in community-dwelling older adults (external test set)

| Variables | Model 1 (age- and sex-adjusted) |  | Model 2 (model 1+ prevalent vertebral fracture and clinical biomarkers) |  |
| --- | --- | --- | --- | --- |
|  | Adjusted HR (95% CI) | P-value | Adjusted HR (95% CI) | P-value |
| Predicted age difference (spine age minus chronological age), per 1 SD increment | 1.32 (1.13 to 1.54) | <0.001 | 1.31 (1.12 to 1.53) | 0.001 |
| Chronological age, per 1 SD increment | 1.93 (1.68 to 2.21) | <0.001 | 1.74 (1.51 to 2.01) | <0.001 |
| Women (vs. men) | 0.28 (0.21 to 0.38) | <0.001 | 0.25 (0.17 to 0.35) | <0.001 |
| Prevalent vertebral fracture |  |  | 1.49 (1.02 to 2.19) | 0.042 |
| Serum albumin, per 1 SD decrement |  |  | 1.18 (1.02 to 1.36) | 0.024 |
| Hemoglobin, per 1 SD decrement |  |  | 1.21 (1.04 to 1.41) | 0.012 |
| Serum creatinine, per 1 SD increment |  |  | 1.12 (1.07 to 1.18) | <0.001 |

Abbreviations: SD, standard deviation, CI, confidence interval.

## Discussion

In this study, spine age estimated from lateral spine radiographs and DXA VFA images using convolutional neural network outperformed chronological age for discriminating presence of morphologic vertebral fracture or osteoporosis in older adults. About 40% of the variance in predicted age difference, calculated as spine age minus chronological age, was explained by chronological age, sex, height, femoral neck BMD, presence of diabetes mellitus, morphologic vertebral fracture, and surgical prosthesis in the spine images. Higher predicted age difference was associated with greater risk of fracture and mortality independent of prevalent vertebral fracture, osteoporosis, and other covariates. Utilizing predicted spine age instead of chronological age to calculate FRAX probabilities yielded modest improvements in the discriminatory performance for incident fracture events.

Several studies have shown the potential of artificial intelligence and machine learning techniques to enhance fracture risk assessment, mostly by improving detection of prevalent vertebral fracture or osteoporosis in plain radiographs or computed tomography images.^7–9^ In a study of longitudinal DXA whole body images to predict mortality, features derived from the raw whole body DXA images using deep learning predicted all-cause mortality with and without clinical risk factors.^10^ Recurrent neural network models using sequential whole body DXA scans outperformed the comparable model using only one observation. This study indicated that deep learning models could be trained to capture information on what constitutes healthy aging from simple two-dimensional musculoskeletal images, beyond the detection of known risk factors such as prevalent fracture or osteoporosis. In line with this notion, we confirmed the feasibility of estimating biological spine age from simple imaging sources such as lateral spine radiographs or DXA VFA by training a deep learning model. Predicted age difference was associated not only with incident fracture outcomes but also with mortality, indicating the potential utility of estimated spine age as an image-derived biomarker for aging-related outcomes beyond fracture risk assessment.

Factors contributing to the difference between estimated spine age and chronological age were identified from a multivariable linear regression model. We observed an association of male sex with accelerated spine aging, along with known risk factors such as lower height^11^, lower femoral neck BMD^12^, presence of diabetes mellitus^13^, morphologic vertebral fracture^14^, and surgical prosthesis^15^ in spine images. A study of opposite-sex twins reported that men were biologically older than women and the association of sex with accelerated aging was stronger in older twins using epigenetic clocks, which supports sex disparity observed in spine age.^16^ Despite the inclusion of well-established risk factors for fracture, the model only explained 40% of variance in predicted age difference, indicating that some information in the spine images was not considered in the explainability model but contributed substantially to spine age estimation. As highlighted in GRAD-CAM images, presence of aortic calcification could be associated with accelerated spine aging.^17^ Degenerative change in the spine could also contribute to estimated spine age.^18^ The association of predicted spine age difference with incident fracture and mortality might be partly mediated by central obesity and low lean mass which affect soft tissue in spine radiographs, though this needs to be explored in future studies. Taken together, these findings portend that biological age estimated from lateral spine radiographs and DXA VFA images have potential to serve as an integrated biomarker of aging and age-related health outcomes.

Estimation of biological age from clinical data sources, including images, may provide new opportunities to improve clinical practice. To test whether biological spine age estimated from lateral spine radiographs and DXA VFA could be useful to improve fracture risk assessment over chronological age, we compared the discriminatory performance of FRAX probabilities for major osteoporotic fracture and hip fracture calculated from both spine age and chronological age. FRAX probabilities based on spine age showed improvement in discriminatory performance for incident fracture over FRAX probabilities based on chronological age. Although the improvement was modest, this finding provides a proof-of-concept example for enhanced fracture risk assessment from inductive, bottom-up deep learning, such as estimated spine age, as a complement to well-established statistical modelling based on hypothesis-driven, top-down domain-specific knowledge^7^.

This study has several limitations. Derivation and test datasets are limited to Korean ethnicities; whether this finding would be applicable to individuals of other ethnicities needs to be examined further. Individuals with ages younger than 40 years were not included in the training dataset. FRAX probabilities were calculated without BMD because BMD data were limited to a subset of study participants in the derivation set. Antero-posterior view spine radiographs were not utilized to calculate spine age; whether utilizing both lateral and antero-posterior view spine images could improve the prediction of spine age merits further investigation. Although estimated spine age from VFA images using model trained in lateral spine radiographs showed similar predictive performance for incident fracture outcome, the correlation of spine age with chronological age was weaker in the VFA cohort. This is partly due to domain shift or reflecting true pattern in older individuals. VFA images from different manufacturers such as GE have different characteristics from VFA images from Hologic and lateral spine radiographs, requiring additional validation. Frailty and physical performance measurements were not available in this study; whether spine age is associated with frailty needs to be studied.

To summarize, spine age estimated from lateral spine radiographs and DXA VFA predicted incident fracture and mortality in adults independent of age, sex, prevalent vertebral fracture, osteoporosis, and other covariates. Spine age outperformed chronological age in discriminating prevalent vertebral fractures and osteoporosis. The discriminatory performance of FRAX probabilities for incident fracture outcome improved modestly when spine age was used as an input variable instead of chronological age.

## Data Availability

Data sharing requests will be considered for the research purpose upon a written request to the corresponding author. If agreed, deidentified participant data and/or deep learning model weights will be made available, subject to a data sharing agreement.

https://github.com/nkhong84/YUHS-VERTE-X-Age

## Acknowledgments

We appreciate the effort of the participants and investigators of the KURE cohort study.

## Author contributions

Sang Wouk Cho: software, methodology, formal analysis, investigation, writing-original draft, visualization, writing-review and editing; Namki Hong: conceptualization, methodology, formal analysis, investigation, data curation, writing-original draft, visualization, writing-review and editing; Kyoung Min Kim: conceptualization, project administration, funding acquisition, investigation, resources, methodology, writing-review and editing; Young Han Lee: writing-review and editing, visualization, methodology; Chang Oh Kim: writing-review and editing, resources, data curation; Hyeon Chang Kim: writing-review and editing, resources, data curation; Yumie Rhee: writing-review and editing, supervision, resources, data curation; Brian H. Chen: methodology, data curation, writing-review and editing; William D. Leslie: methodology, formal analysis, investigation, data curation, writing-review and editing; Steven R. Cummings: conceptualization, supervision, writing-review and editing, supervision; and. All authors have read and finally agreed to the published version of the manuscript.

## Data Sharing Statement

All source codes to train deep learning model to estimate spine age is publicly available on GitHub (https://github.com/nkhong84/YUHS-VERTE-X-Age). The developed models can be tested at the website (https://bonecentriq.org). Data sharing requests will be considered for the research purpose upon a written request to the corresponding author. If agreed, deidentified participant data and/or deep learning model weights will be made available, subject to a data sharing agreement.

## References

1. GBD2019. Global, regional, and national burden of bone fractures in 204 countries and territories, 1990-2019: a systematic analysis from the Global Burden of Disease Study 2019. The lancet Healthy longevity 2021; 2(9): e580–e92.

2. Nguyen TV. Individualized fracture risk assessment: State-of-the-art and room for improvement. Osteoporosis and sarcopenia 2018; 4(1): 2–10.

3. Zhang Q. An interpretable biological age. The lancet Healthy longevity 2023; 4(12): e662–e3.

4. Wood DA, Kafiabadi S, Busaidi AA, et al. Accurate brain-age models for routine clinical MRI examinations. NeuroImage 2022; 249: 118871.

5. Nusinovici S, Rim TH, Li H, et al. Application of a deep-learning marker for morbidity and mortality prediction derived from retinal photographs: a cohort development and validation study. The lancet Healthy longevity 2024; 5(10): 100593.

6. Raghu VK, Weiss J, Hoffmann U, Aerts H, Lu MT. Deep Learning to Estimate Biological Age From Chest Radiographs. JACC Cardiovascular imaging 2021; 14(11): 2226–36.

7. Hong N, Whittier DE, Glüer C-C, Leslie WD. The potential role for artificial intelligence in fracture risk prediction. The Lancet Diabetes & Endocrinology 2024; 12(8): 596–600.

8. Hong N, Cho SW, Shin S, et al. Deep-Learning-Based Detection of Vertebral Fracture and Osteoporosis Using Lateral Spine X-Ray Radiography. J Bone Miner Res 2023; 38(6): 887–95.

9. Smets J, Shevroja E, Hügle T, Leslie WD, Hans D. Machine Learning Solutions for Osteoporosis—A Review. Journal of Bone and Mineral Research 2021; 36(5): 833–51.

10. Glaser Y, Shepherd J, Leong L, et al. Deep learning predicts all-cause mortality from longitudinal total-body DXA imaging. Communications medicine 2022; 2: 102.

11. Lee C, Park HS, Rhee Y, Hong N. Age-Dependent Association of Height Loss with Incident Fracture Risk in Postmenopausal Korean Women. Endocrinol Metab (Seoul) 2023; 38(6): 669–78.

12. Cummings SR, Browner W, Cummings SR, et al. Bone density at various sites for prediction of hip fractures. The Lancet 1993; 341(8837): 72–5.

13. Zoulakis M, Johansson L, Litsne H, Axelsson K, Lorentzon M. Type 2 Diabetes and Fracture Risk in Older Women. JAMA Network Open 2024; 7(8): e2425106-e.

14. Lentle BC, Berger C, Probyn L, et al. Comparative Analysis of the Radiology of Osteoporotic Vertebral Fractures in Women and Men: Cross-Sectional and Longitudinal Observations from the Canadian Multicentre Osteoporosis Study (CaMos). Journal of Bone and Mineral Research 2018; 33(4): 569–79.

15. Nakahashi M, Uei H, Tokuhashi Y, et al. Vertebral fracture in elderly female patients after posterior fusion with pedicle screw fixation for degenerative lumbar pathology: a retrospective cohort study. BMC Musculoskeletal Disorders 2019; 20(1): 259.

16. Kankaanpää A, Tolvanen A, Saikkonen P, et al. Do Epigenetic Clocks Provide Explanations for Sex Differences in Life Span? A Cross-Sectional Twin Study. The Journals of Gerontology: Series A 2022; 77(9): 1898–906.

17. Gebre AK, Lewis JR, Leow K, et al. Abdominal Aortic Calcification, Bone Mineral Density, and Fractures: A Systematic Review and Meta-analysis of Observational Studies. The journals of gerontology Series A, Biological sciences and medical sciences 2023; 78(7): 1147–54.

18. Ye C, Leslie WD, Bouxsein ML, et al. Association of vertebral fractures with worsening degenerative changes of the spine: a longitudinal study. J Bone Miner Res 2024.

19. Hong N, Kim KJ, Lee SJ, et al. Cohort profile: Korean Urban Rural Elderly (KURE) study, a prospective cohort on ageing and health in Korea. BMJ Open 2019; 9(10): e031018.

20. Lee EY, Kim HC, Rhee Y, et al. The Korean urban rural elderly cohort study: study design and protocol. BMC geriatrics 2014; 14: 33.

21. Pan H, Han H, Shan S, Chen X. Mean-Variance Loss for Deep Age Estimation from a Face. 2018 IEEE/CVF Conference on Computer Vision and Pattern Recognition; 2018 18-23 June 2018; 2018. p. 5285–94.

22. Zhang B, Zhang S, Feng J, Zhang S. Age-level bias correction in brain age prediction. NeuroImage Clinical 2023; 37: 103319.

23. Beheshti I, Nugent S, Potvin O, Duchesne S. Bias-adjustment in neuroimaging-based brain age frameworks: A robust scheme. NeuroImage Clinical 2019; 24: 102063.

24. Selvaraju RR, Cogswell M, Das A, Vedantam R, Parikh D, Batra D. Grad-CAM: Visual Explanations from Deep Networks via Gradient-Based Localization. 2017 IEEE International Conference on Computer Vision (ICCV); 2017 22-29 Oct. 2017; 2017. p. 618–26.

25. Kim JA, Yoon S, Kim LY, Kim DS. Towards Actualizing the Value Potential of Korea Health Insurance Review and Assessment (HIRA) Data as a Resource for Health Research: Strengths, Limitations, Applications, and Strategies for Optimal Use of HIRA Data. J Korean Med Sci 2017; 32(5): 718–28.

26. Cosman F, de Beur SJ, LeBoff MS, et al. Clinician’s Guide to Prevention and Treatment of Osteoporosis. Osteoporos Int 2014; 25(10): 2359–81.

27. Looker AC, Orwoll ES, Johnston CC, Jr., et al. Prevalence of low femoral bone density in older U.S. adults from NHANES III. J Bone Miner Res 1997; 12(11): 1761–8.

28. DeLong ER, DeLong DM, Clarke-Pearson DL. Comparing the areas under two or more correlated receiver operating characteristic curves: a nonparametric approach. Biometrics 1988; 44(3): 837–45.

