## Supplementary for "Spine age estimation using deep learning in lateral spine radiographs and DXA VFA to predict incident fracture and mortality"

**Supplementary file**

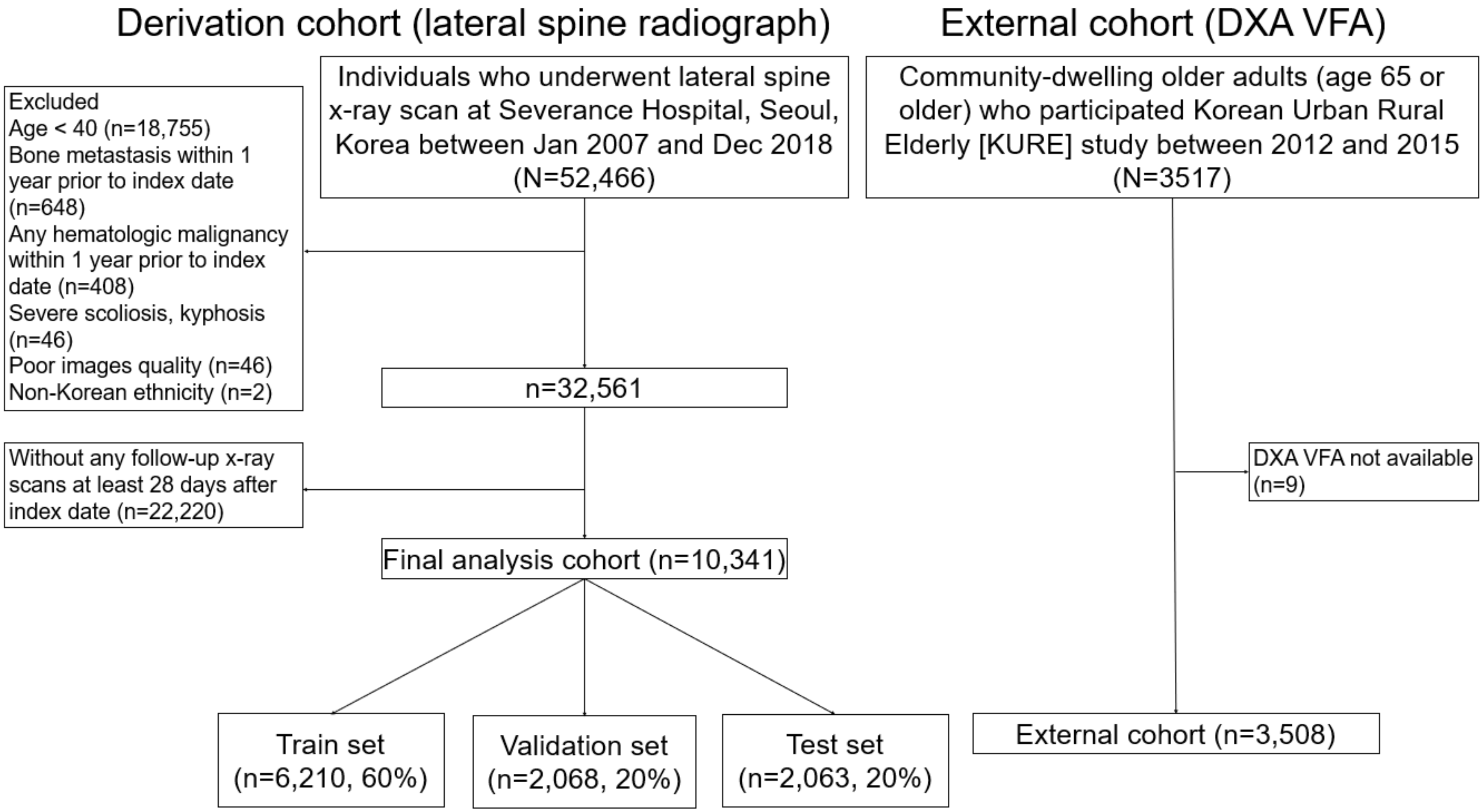


Supplementary Figure 1. Study flow

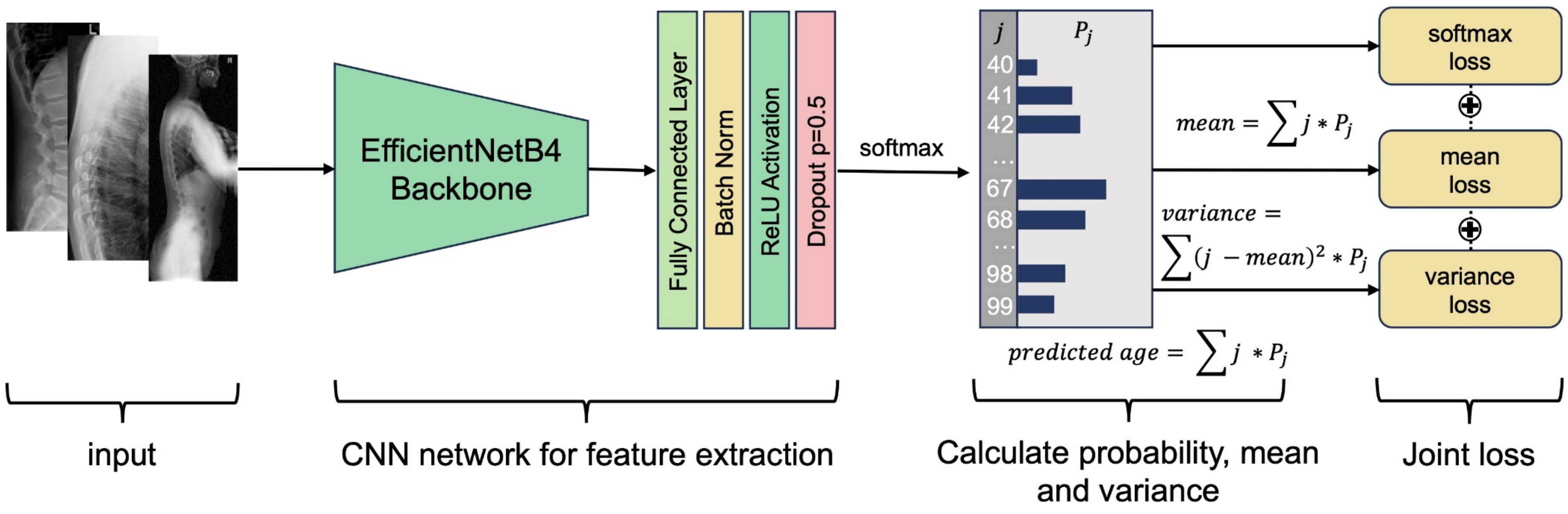


Supplementary Figure 2. Deep learning model architecture to predict spine age

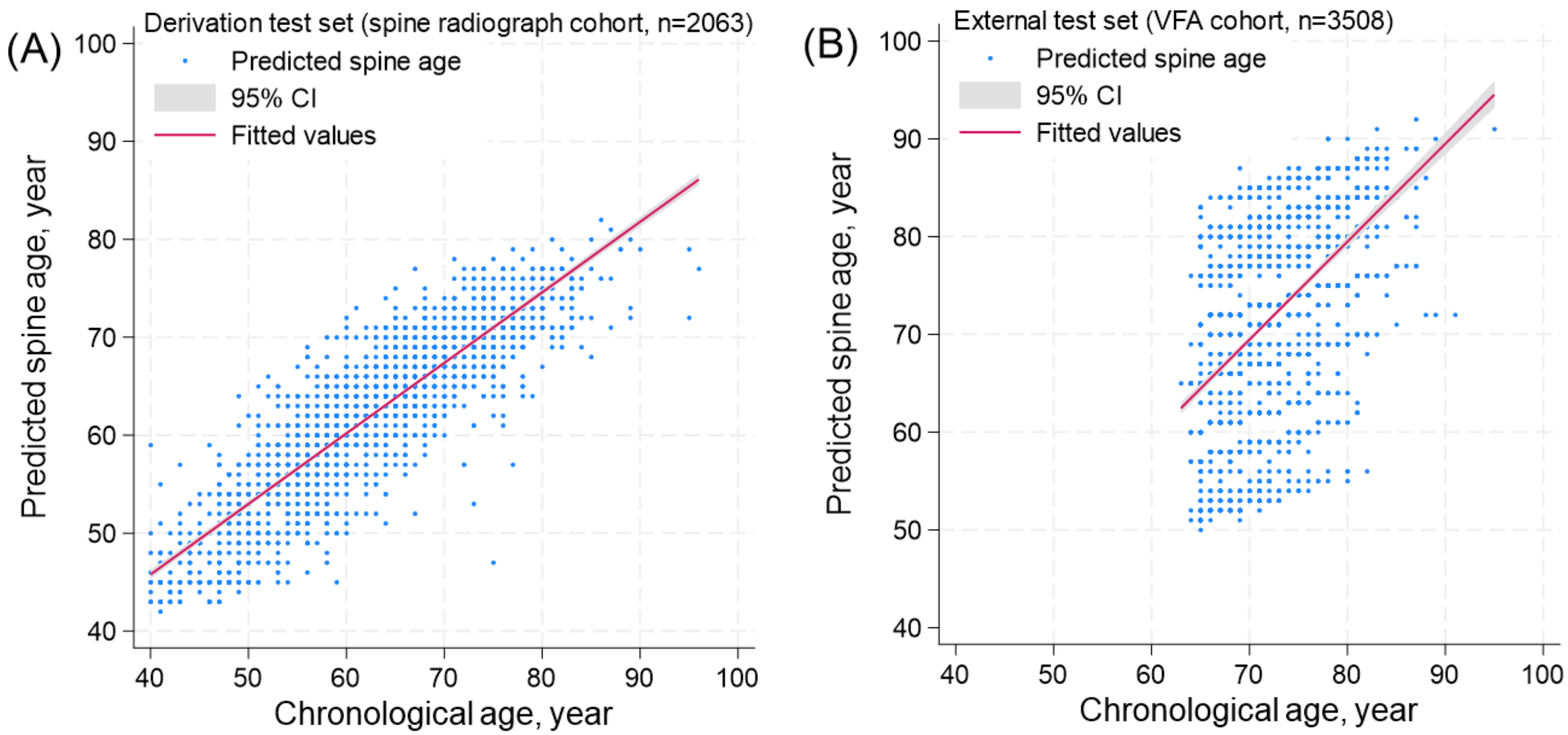


Supplementary Figure 3. Associations of chronological age and predicted spine age in the (A) derivation test set (spine radiograph cohort, aged 40 years or older n=2063) and (B) external test set (DXA VFA cohort, age 65 years or older, n=3508).

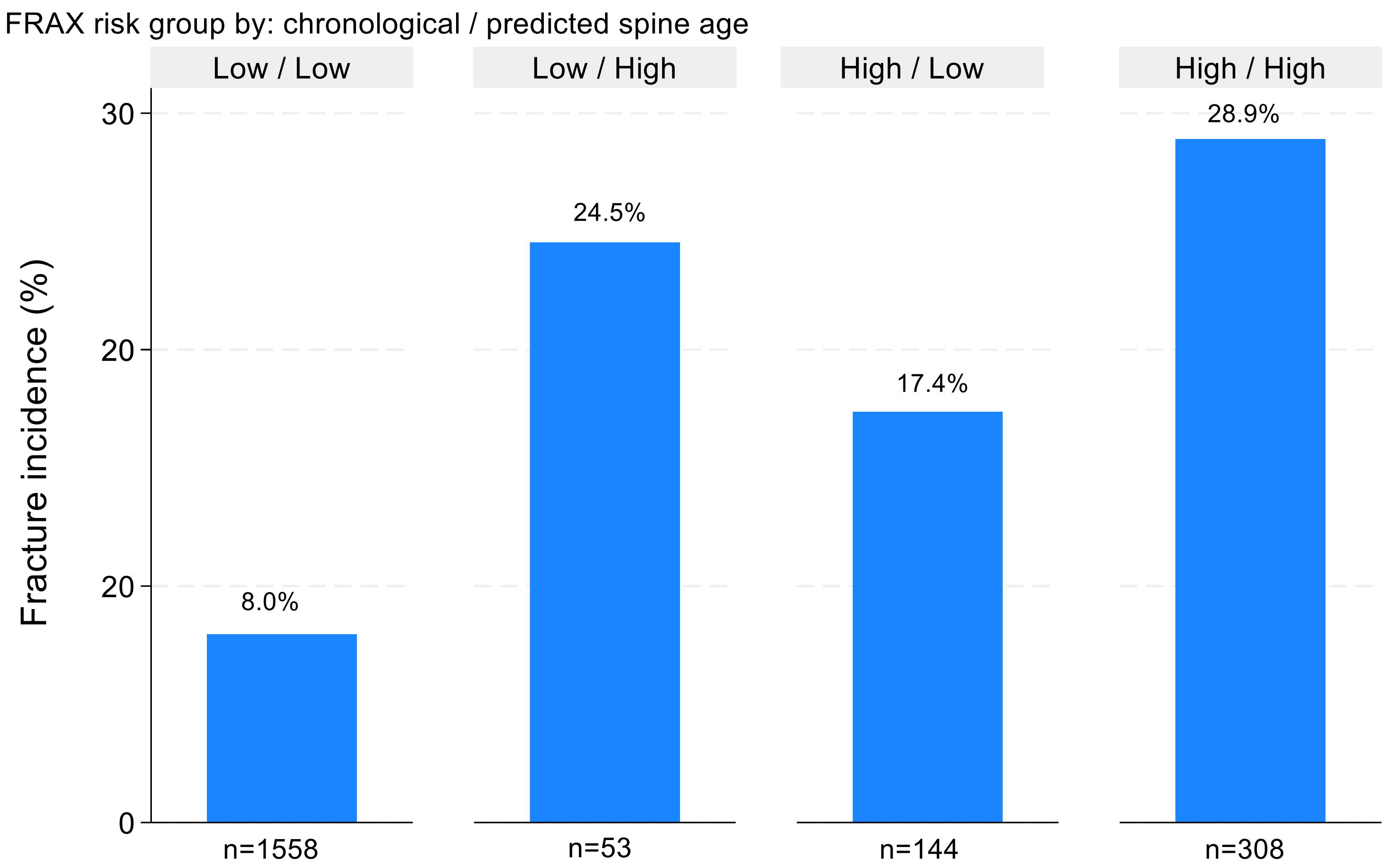


Supplementary Figure 4. Incidence of overall fracture by FRAX risk groups in lateral spine radiograph cohort test set (n=2063). High risk group was defined as FRAX major osteoporotic fracture probability >20% or hip fracture probability >3%. FRAX probability groups calculated using chronological age (high and low) were reclassified by FRAX recalculated using predicted spine age, creating a total of four groups. Low/High group indicates upward reclassification to high risk by FRAX using predicted spine age (n=53), whereas High/Low group indicates downward reclassification to low risk (n=144).

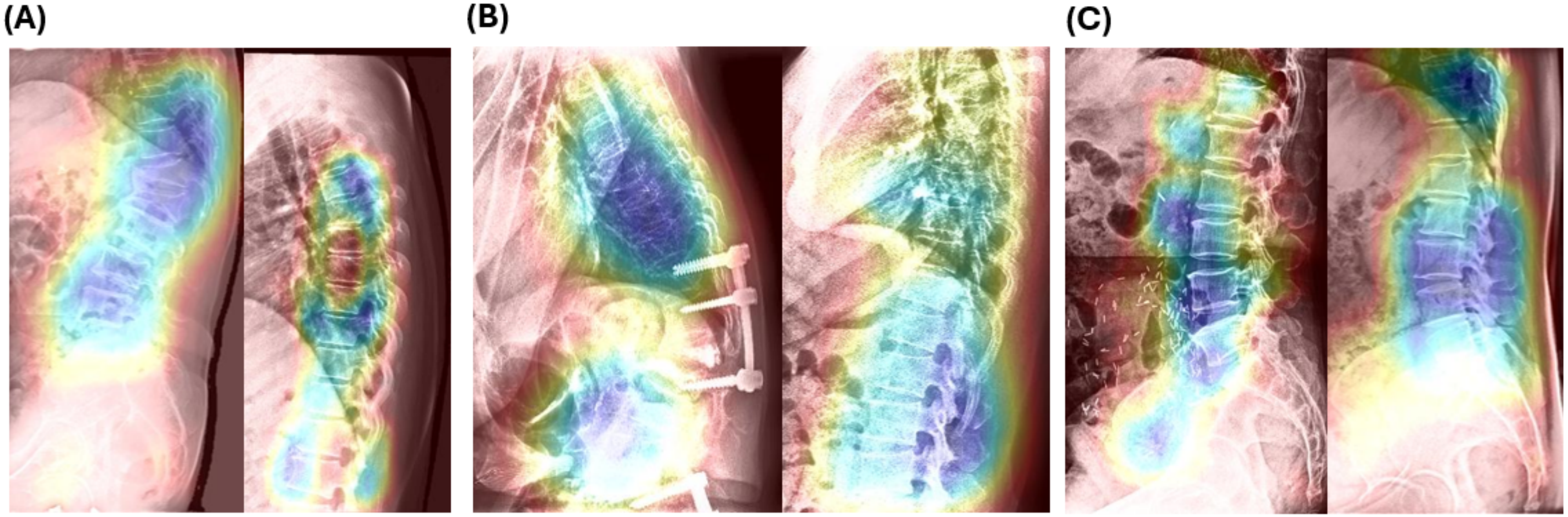


| | | | | | | |
|---|---|---|---|---|---|---|
| Chronologic age (year) | 66 | 63 | 60 | 60 | 58 | 58 |
| Predicted spine age (year) | 70 | 59 | 70 | 60 | 67 | 58 |

Supplementary Figure 5. Examples of gradient-weighted class activation map (GRAD-CAM) of lateral spine radiographs to predict spine age. Left panels in figure 4A, 4B, and 4C represent images with accelerated spine age, whereas right panels represent images without accelerated spine age. Examples with accelerated spine age show (A) presence of morphologic vertebral fracture, (B) vertebroplasty, surgical prosthesis, (C) and aortic calcification, with various degrees of degenerative changes.

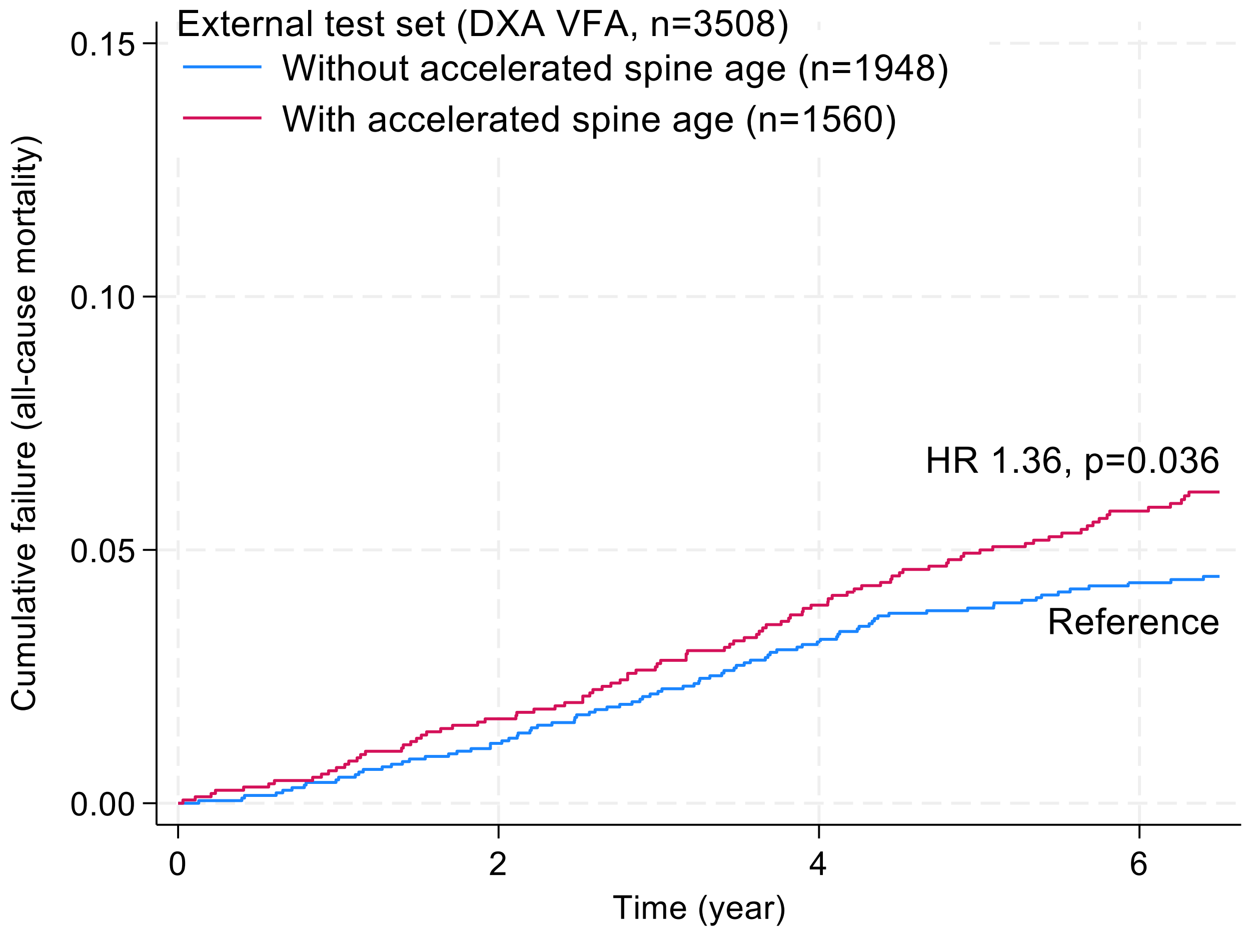


Supplementary Figure 6. Kaplan-Meier curve for association between accelerated spine age and all-cause mortality in community-dwelling older adults (external test set, DXA VFA). Accelerated spine age was defined as the highest tertile group of predicted age difference (spine age – chronological age, +1 or higher [years]) observed in the derivation test set.

Supplementary Table 1. Performance of FRAX probability based on chronological age and predicted spine age to discriminate incident fracture events in lateral spine radiograph cohort

| High risk vs. low risk groups (High risk: FRAX MOF risk ≥ 20% or hip fracture risk ≥ 3%) | FRAX probability estimated using chronological age | FRAX probability estimated using predicted spine age |
|---|---|---|
| Prevalence, % | 21.9 | 17.5 |
| Sensitivity, % | 45.4 | 40.6 |
| Specificity, % | 81.3 | 85.7 |
| Positive predictive value, % | 25.2 | 28.3 |
| Negative predictive value, % | 91.5 | 91.2 |
| Odds ratio | 3.6 | 4.1 |

Abbreviations: MOF, major osteoporotic fracture.

Supplementary Table 2. Discriminatory ability of FRAX probability based on chronological age and predicted spine age for incident fractures in lateral spine radiograph cohort

| | AUROC (95% CI) | | P-value |
|---|---|---|---|
| Outcomes | FRAX MOF probability* (chronological age) | FRAX MOF probability (predicted spine age) | |
| Incident overall fracture | 0.73 (0.69 to 0.76) | 0.74 (0.71 to 0.77) | 0.097 |
| Incident hip fracture | 0.77 (0.72 to 0.82) | 0.81 (0.76 to 0.86) | 0.007 |
| | FRAX HF probability (chronological age) | FRAX HF probability (predicted spine age) | |
| Incident overall fracture | 0.72 (0.69 to 0.75) | 0.74 (0.71 to 0.77) | 0.024 |
| Incident hip fracture | 0.78 (0.73 to 0.84) | 0.83 (0.77 to 0.87) | 0.027 |

* FRAX probabilities without BMD. Abbreviations: MOF, major osteoporotic fracture; HF, hip fracture.

Supplementary Table 3. Factors associated with predicted age differences in the subset of the derivation test set (spine radiograph cohort, n=1,448) with available DXA BMD data

| Variables | Change in predicted age difference, adjusted (95% CI) | P-value |
| --- | --- | --- |
| Age, per 1 year increment | -0.34 (-0.36 to -0.31) | <0.001 |
| Women (vs. men) | -0.66 (-1.29 to -0.02) | 0.043 |
| Height, per 5 cm decrement | +0.31 (+0.13 to +0.49) | 0.001 |
| Presence of diabetes | +0.51 (+0.05 to +0.97) | 0.031 |
| Prevalent morphometric vertebral fracture | +1.64 (+1.06 to +2.21) | <0.001 |
| Femoral neck BMD, per 1 SD decrement | +0.64 (+0.40 to +0.86) | <0.001 |
| Surgical prosthesis in spine | +1.15 (+0.51 to +1.80) | <0.001 |

Adjusted $R^2$ of the multivariable model: 0.40

### Supplementary Methods

#### *1. Image processing*

Because of the intensity difference in individual images, histogram equalization was applied to all images, and Min-Max scaling method was selected to normalize the pixel intensity values of the images. About 5% of the image size was cropped for areas not related to analysis, and the cropped images were resized according to the size (1024, 512). Since the width and height differed by each image, we set the width to 512 px if the width was larger than the height, then resized the height according to the resolution ratio. If an image was smaller than (1024,512), the image was aligned to the center position with zero-padding to the rest of the area. As VFA images had a narrower lateral width with focus on the thoracic and lumbar spine, no cropping was needed for the VFA images.

#### *2. Mean-variance loss used in the spine age prediction model*

While other studies addressing age prediction used exact age regression models, we found that exact regression does not effectively leverage the robustness of distributions in capturing labels with inherent ambiguity. Given that both X-ray and VFA images are grayscale, limiting their expressiveness and diversity compared to RGB images, it was essential to incorporate distributional analysis. To address this, we applied the mean-variance loss function proposed by Pan (2019).

#### *3. Deep learning model architecture*

Our deep learning models predicted the spine age by receiving spine X-ray images and VFA images. It was built based on the Efficientnet-b4 model using Adam optimizer with the initial learning rate of 1e-4. To the prevent overfitting, weight decay was applied to the models. Due to the size of spine X-ray images and memory limitations, the batch size was set to 45. The experiments were up to 100 epochs on 4 NVIDIA RTX 3090 Graphic processor units (GPU). After each training epoch, the models were evaluated by Mean squared errors (MSE) and loss of validation set. There were 1000 logit values of ImageNet in last layer of original Efficient-net b4 model, but we modified the number of fully connected (FC) layer which concatenated with FC layer of ages in our models. The final output is a value with 60 layers (Age 40~100), which are then used to compute cross-entropy loss ($L_c$), mean loss ($L_m$), and variance loss ($L_v$), which together drive model training through backpropagation.

The mean loss penalizes the difference between the mean of an estimated spine age distribution and the ground-truth age. Different from softmax loss which focuses on classification tasks, our mean loss emphasizes on regression tasks, and we use the L2 distance to measure the distance between the mean of an estimated age distribution and the ground-truth age. Therefore, it is complementary to the softmax loss. Such a variance loss requires that an estimated distribution should be concentrated at a small range of the mean. The variance loss penalizes the dispersion of an estimated spine age distribution, making it as sharp as possible. This is helpful to obtain an accurate spine age estimation with a narrow confidence interval.

We applied this mean-variance loss into the architecture of convolutional neural network model, and the softmax loss and mean-variance loss was used jointly as the supervision signal. The final Loss of the spine age prediction model could be represented as:

$$L_{final} = L_c + \lambda_1 L_m + \lambda_2 L_v$$

where $\lambda_1$ and $\lambda_2$ are two hyper-parameters, balancing the influencing of individual sub-losses in the joint loss. Initially, $\lambda_1$ and $\lambda_2$ was 0.2 and 0.05, respectively.

In the inference phase, the age of a test image is estimated as:

$$y_p = r(\sum_{i=40}^{K} i * p_i)$$

where $p_i$, $i \in \{40,41,42,\ldots,K\}$ is the output of the softmax layer in the network, and $r(\cdot)$ is a round function.
